# Clinicians’ perspective on use of immune checkpoint inhibitors and related biomarkers for solid tumors

**DOI:** 10.1101/2022.04.01.22269254

**Authors:** Kaitlynn Moser, Michelle Soto, Daniela Geba, Paul Allen, Klaus Lindpaintner

## Abstract

**Background:** Immune checkpoint inhibitors (ICIs) are a valuable treatment option for patients with malignant tumors, but only selected patients respond to ICIs. Available biomarkers are of limited use in guiding ICI therapy.

**Objective:** To examine clinicians’ perspective on the use of ICIs and biomarkers for treatment of malignant tumors and to identify unmet needs related to their use.

**Methods:** We conducted in-depth telephone interviews of eight oncologists, and 100 oncologists completed online surveys.

**Results:** Oncologists have a positive attitude toward use of ICIs, and 98% of them prescribe them in all approved indications. Clinicians report that only about half of the patients with solid tumors responded to treatment, overestimated the response rate to ICIs across most types of tumors they treat compared with data in the literature. They ranked the lack of reliability of biomarkers to guide treatment (rating of 4.4 out of 7) as the top challenge with use of ICIs, followed by lack of overall efficacy and toxicity or occurrence of immune-related adverse events. The biomarkers most often used by survey participants were: a comprehensive panel including driver mutations and tumor mutational burden(69% of respondents), programmed cell death ligand-1 (PD-L1) expression (62%), and microsatellite instability (MSI) (56%). Oncologists indicated that they ordered biomarkers for each type of cancer according to their perceived usefulness of each biomarker in predicting the outcomes for ICI therapy, being more likely to use those perceived as useful or very useful.

**Conclusion:** Clinicians indicate that more reliable therapy-response prediction biomarkers would have a great impact on treatment decisions for patients with solid tumors, reducing unnecessary treatments, side effects, and health care expenditures.

## INTRODUCTION

Immunotherapy has emerged as a revolutionary new therapeutic approach in the field of oncology and has become a cornerstone in cancer therapy, with immune-check-point inhibitors (ICIs) currently being the most commonly used type of immunotherapy^1^. Over the past decade, this type of systemic cancer therapy has been integrated in the therapeutic pathways of a number of solid and hematologic malignancies, such as non-small cell lung cancer (NSCLC), malignant melanoma, head and neck cancer, urothelial and renal cell carcinoma (RCC), gastrointestinal cancer, and lymphoma^2,3^. The number of ICIs approved by regulatory agencies has continued to grow in recent years and currently includes the following classes and drugs: programmed cell death protein 1 (PD-1) inhibitors (nivolumab, pembrolizumab, pidilizumab, and cemiplimab), programmed cell death ligand-1 (PD-L1) inhibitors (atezolizumab, durvalumab, and avelumab), and cytotoxic T-lymphocyte-associated antigen-4 (CTLA-4) inhibitors (ipilimumab and tremelimumab). ICIs may be used as monotherapy or in combination, and as first-line, second-line, adjuvant, and neo-adjuvant therapy^3^.

Unlike traditional cytotoxic chemotherapy, ICIs enhance the body’s immune system’s antitumor activity, effecting tumor regression even in in patients with advanced or end-stage cancer and inducing and augmenting immunological memory that can confer long-term protection against cancer recurrence. However, current experience shows that only a fraction of patients achieve long-term response with ICIs^4^. The response rate to ICIs varies significantly by type of cancer. While for some types of cancer, such as melanoma and Hodgkin’s lymphoma, studies report response rates for treatment with PD-1 inhibitors ranging from 40% to 70%^5-7^, for other diseases, such as NSCLC, urothelial carcinoma, and RCC, the response rate is reported in the 10% to 25% range^8-12^. Over time, the response rate is reduced even further, as disease progression can develop among patients who initially respond to ICIs but later become resistant to ICIs^13^. Administration of immunotherapy is frequently associated with immune-related adverse effects (irAEs) which can be severe^14^. An additional limitation of ICIs is the high financial burden on health care systems, which relates to the cost of the ICIs and concomitant treatments^15^.

A growing number of biomarkers, such as PD-L1 expression, nuclear protein Ki-67 expression, tumor mutational burden (TMB), and microsatellite instability (MSI), and cell-based biomarkers, such as tumor infiltrating lymphocytes and myeloid derived suppressor cells, are currently used for predicting the outcome of ICI therapy^16-18^. Because of their limited accuracy in predicting therapy response, none of these biomarkers have proven to be truly helpful in guiding ICI treatment choices. In a meta-analysis of all FDA approvals for ICIs from 2011 to 2019 across 15 tumor types, PD-L1 was predictive in 29% of the approvals, was not predictive in 53% of the cases, and was not tested in the remaining cases (18%)^19^. The results of another meta-analysis showed a sensitivity of 64% and specificity of 49% for PD-L1 expression level across all cancer type based on results of 76 studies (aggregate area under the receiver operating characteristic curve (AUC) of 0.58) and a sensitivity of 70% and specificity of 53% for TMB across all cancer type based on results of 15 studies (AUC of 0.65)^20^. Further, the AUCs were inconsistent across biomarkers with regard to cancer type, as shown, for example, for TMB, which has a sensitivity of 58%, specificity of 69%, and an AUC of 0.7, respectively, for NSCLC and a sensitivity of 86%, specificity of 36%, and an AUC of 0.37, respectively, for melanoma.

Recently, a novel biomarker platform based on accessing and interrogating specific glycosylation patterns of peripheral blood proteins using mass spectrometry coupled to artificial intelligence-based data processing has been found to predict therapy response to ICIs with superior performance as compared to currently available biomarkers in several solid cancers, such as NSCLC^21^. Availability of these more accurate predictive biomarkers for ICI therapy is expected to have significant impact on the treatment for patients with tumors for which ICIs have been approved, by allowing a more targeted selection of patients likely to benefit from ICI treatment while avoiding unnecessarily exposing likely non-responders to ICI-related adverse effects as well as reducing the overall cost of ICI prescriptions.

Given the growing importance of ICI treatment and of predictive biomarkers guiding oncologists in their choice of treatment options, we conducted a study which included in-depth interviews and two online surveys among oncologists to examine clinicians’ perspective on the use of ICIs and biomarkers for ICI treatment for solid tumors and to identify currently unmet needs related to their use.

## METHODS

### Study design and participants

The study included a descriptive qualitative phase and a subsequent quantitative phase. The study participants were recruited from an online market research panel. The qualitative phase was conducted in February 2021 and consisted of 60-minute, web-assisted semi-structured telephone interviews of eight board-certified oncologists with four key opinion leaders (KOLs) and four non-KOLs. KOLs were defined as clinicians who met at least two of the three criteria: they spent at least 75% of their professional time at a teaching hospital that is a member of the National Comprehensive Care Network (NCCN) or National Cancer Institute (NCI), they had acted as a principal investigator or sub-investigator for clinical studies on ICIs treatments or ICI-related biomarkers, or they had presented ICI-related research results at a national or regional conference. Requirements for participation in the interview were: specialty board certification in oncology, clinical experience between 3 and 30 years, proportion of total professional time devoted to direct patient care of at least 50% for those with academic affiliation and at least 70% for those in community practice settings, and at least half of the time spent treating patients with solid tumors for non-KOLs. Recruitment was designed to ensure an equal representation of oncologists from academic centers and community practices across the medical oncology-hematology specialties. The scope of the interviews was to examine the respondents’ perceptions of ICIs, of currently available biomarker testing, and of the need for novel blood-based tests to predict response to ICIs. With the permission of the participants, in-depth interviews were recorded and transcribed verbatim after the interview. Data collected during the qualitative phase were used to design the questionnaires for the quantitative phase.

The quantitative phase included a 15-minute online survey, fielded during three consecutive weeks in April and May 2021, and a 5-minute online survey fielded in September 2021, to address additional questions that had surfaced based on the previous survey. The inclusion criteria and the topics covered by the survey were the same as those used for the qualitative phase. Participants in the first survey were also invited to complete the second survey, which included additional questions on the choice of ICIs for selected solid tumors and on the accuracy of predictive biomarkers for ICI treatment.

### Bioethical considerations, consent, and permissions

The interviews and surveys were conducted in accordance with the principles and guidelines established by the Office for Human Research Protections and the Insights Association Code of Standards and Ethics. The study protocol was reviewed and approved by the Western Institutional Review Board-Copernicus Group (WCG IRB, Pyallup, WA 98374, USA) under protocol number #1-1531933-1. Prior to the start of the research, potential participants were informed about the purpose and nature of the study and that the information would be collected anonymously. Those who consented to participate in the study were admitted to the screening portion of the interview or survey. Respondents could discontinue the interview or survey at any time. Respondents were offered an industry-standard honorarium for their time and effort.

### Statistical analyses

We performed descriptive statistical analyses (means, frequencies) of data collected in the surveys using a MarketSight software. Chi-squared tests were used to compare categorical variables and t-tests were used for comparison of continuous variables. Statistics were unweighted. P values of less than 0.05 were considered statistically significant.

## RESULTS

### Sample characteristics

A total of 108 oncologists participated in the study: eight respondents completed the in-depth interviews, and 100 respondents completed the online survey. The mean length of experience was 26 years in clinical practice for the interview respondents and 17 years for the survey respondents. Three-quarters of the survey respondents practiced in community hospitals and one-quarter had academic affiliations. The average number of patients treated by respondents ranged from an average of 49 patients with melanoma to 100 patients with NSCLC. More than half of respondents perceived themselves as early adopters of new medical products, and only 3% of the sample viewed themselves as late adopters. Selected characteristics of the study participants are summarized in **Table 1**. A total of 70 survey respondents also completed the follow-up survey; their characteristics were not materially different from those of the sample which completed the first survey.

**Table 1.** Characteristics of the study participants.

|  | Qualitative study<br>(n=8) | Quantitative study |  |
| --- | --- | --- | --- |
|  |  | Initial survey<br>(n=100) | Follow-up survey<br>(n=70) |
| **Hospital affiliation** |  |  |  |
| Academic | 4 (50%) | 26 (26%) | 17 (24%) |
| Community | 4 (50%) | 74 (74%) | 53 (76%) |
| **Participant category** |  |  |  |
| KOL | 4 (50%) | 26 (26%) | 16 (23%) |
| Non-KOL | 4 (50%) | 74 (74%) | 54 (77%) |
| **Medical specialty** |  |  |  |
| Medical oncology | 4 (50%) | 37 (37%) | 25 (36%) |
| Hematology and oncology | 4 (50%) | 63 (63%) | 45 (64%) |
| **Years in practice, average (± SD)** | 24 (± 5.2) | 17 (± 6.9) | 17 (± 6.8) |
| **Time spent treating solid tumors**<br>**(% of clinical time)** | 73% | 67% | 69% |

Survey participants were asked about their practice related to the use of ICIs for treatment of solid tumors. Almost all oncologists (98%) reported using ICIs in all approved indications, while only slightly more than one-third of the sample (37%) reported that they had ever used ICIs in non-approved indications (e.g., rare tumors, such as glioblastoma). According to them, most patients with solid tumors received an ICI at some point in their treatment journey, but the percentage varied greatly by type of cancer. In the year prior to the survey, the respondents most often prescribed ICIs as part of first-line combination therapy for NSCLC (43%) and as first-line monotherapy for treatment of melanoma (38%). Overall, gastrointestinal cancers were least often treated with ICIs by the survey respondents during the same period (**Figure 1**).

**Figure 1.**
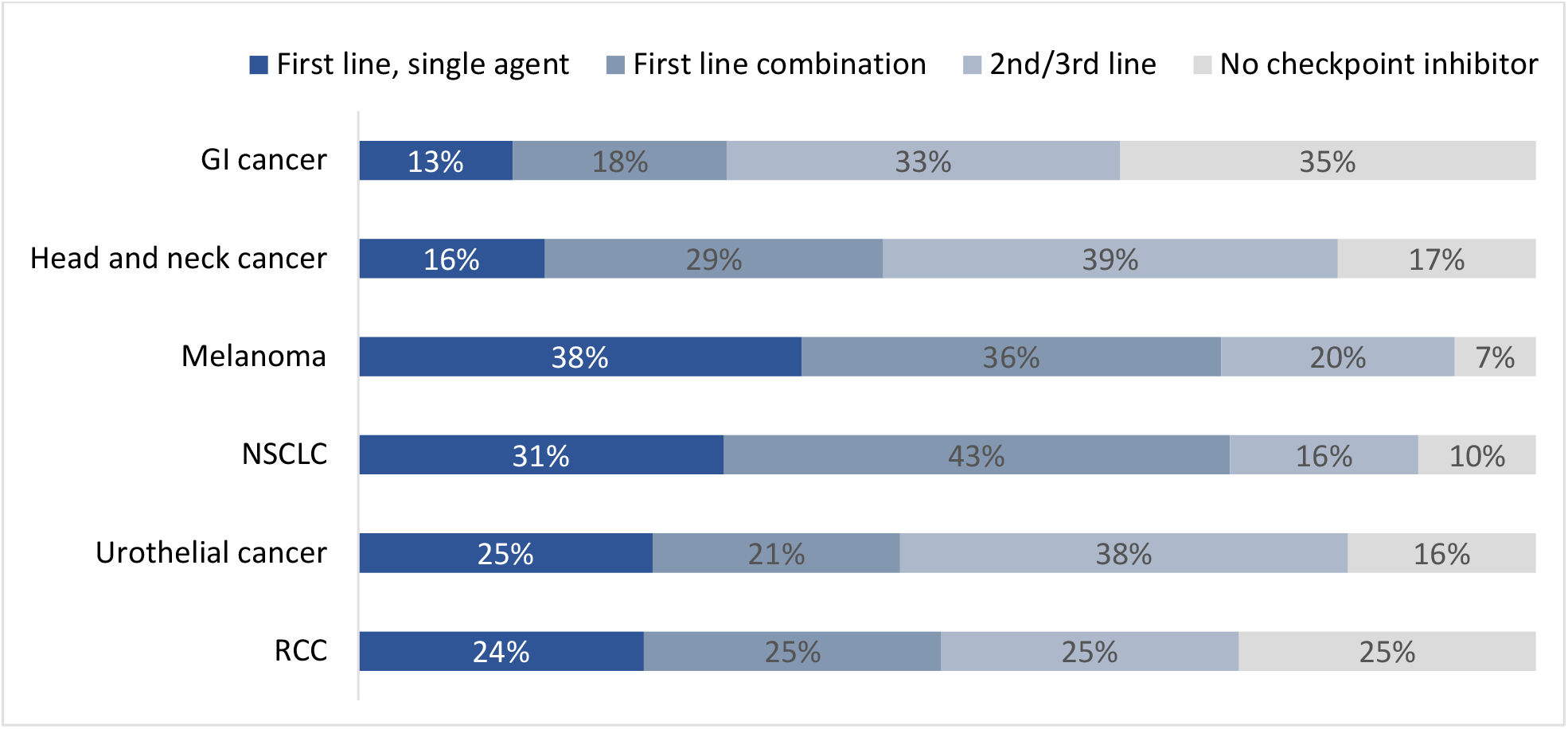
Percentage of patients with solid tumors who were treated by survey respondents and received ICIs during the previous year (n=100 respondents)

The survey respondents indicated that about half of the patients with solid tumors responded to treatment with ICIs, with response rate varying by cancer type. Specifically, melanoma was ranked as having the highest response rate to ICIs (61%), followed by NCCLC (56%) and RCC (56%), whereas gastrointestinal cancers were ranked lowest (43%) (**Figure 2**). Of note, except for gastrointestinal cancers, non-KOLs reported significantly higher response rate to ICIs compared to KOLs, with differences as high as 15% (e.g., for melanoma, reported response rates of 65% and 50%, respectively).

**Figure 2.**
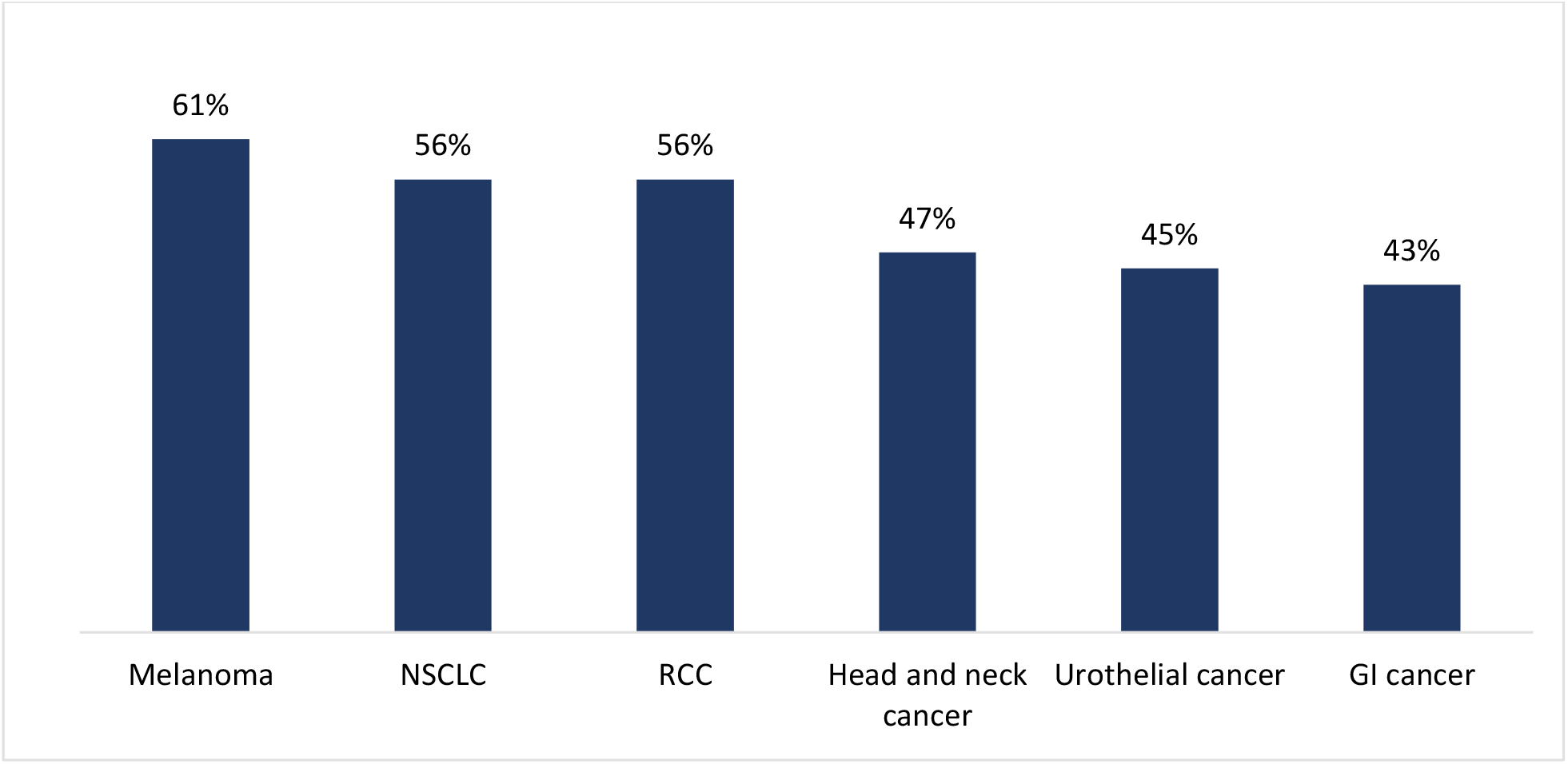
Percentage of patients who were treated with ICIs by respondents during the previous year and responded to treatment (n= 100 respondents) NSCLC: non-small cell lung cancer; RCC: renal cell carcinoma; GI: gastrointestinal

Respondents reported lack of reliability of biomarkers for ICI efficacy prediction as the top challenge, with a mean rating of 4.4 on a 7-points scale (only 24% rating it as a 6 or a 7), whereas the lack of overall efficacy and of irAEs were rated as challenging by only 13% and 9% of the respondents, respectively.

### Perception and use of biomarker testing for prediction of response to ICIs

Interview respondents indicated that biomarker testing was typically done early on in the clinical journey of patients with cancer, if not immediately at the time of biopsy. Testing was viewed as influencing treatment decisions and, for selected types of cancer, it was considered very important for obtaining insurance coverage. Some reported that ordering next generation sequencing of comprehensive biomarker panels had become “reflexive,” whereas other respondents reported that they were not prone to ordering biomarker testing for treatment decisions which were not dependent on biomarker results (e.g., melanoma, RCC, bladder cancer, head and neck cancer).

> “I only do the biomarker testing if it’s going to affect my choice of therapy. And in head and neck cancer, evidently, most of them benefit, whether they have PDL-1 or not. So, then I wouldn’t check it.” – verbatim comment by KOL
>
> “Basically, if it’s not necessary, we don’t do it.” - verbatim comment by KOL

Regarding the type of testing used in their clinical practice, microsatellite instability (MSI) was viewed by respondents as the most predictive biomarker for most tumors. Respondents referenced the need for PD-L1 expression biomarker testing to prescribe drugs, stating that a positive test would establish eligibility for ICI-monotherapy. TMB was regarded as less predictive than the other biomarkers, depending on the cancer type.

Respondents agreed that currently available biomarkers were not sufficiently predictive and that there is a clear need for better biomarkers. They stated that with regard to novel biomarkers they would want to have a comprehensive understanding of the nature of these biomarkers and would want to see the results of studies conducting head-to-head comparisons between them and currently available ones.

> “…the data showed there is a clear proportional response rate to PD-L1 level, but we often see good responses in patients that are PD-L1 negative. It may not be as predictive as hoped; it would be useful to have a marker that better predicts a response to these.” – verbatim comment by Oncologist, Academic hospital

Survey participants were asked about their practice related to biomarker testing for ICI treatment of solid tumors. Two-thirds of respondents indicated that they ordered biomarker testing on tissue samples for patients with solid tumors and only 17% each ordered either testing on blood samples or both tissue and blood samples. A comprehensive panel was most often used among survey participants, as reported by 69% of them, followed by PD-L1 expression (62%) and MSI (56%). Among single tests, PD-L1 expression was tested most often, with almost all (99%) using it for NSCLC, and 82% and 81% testing it for urothelial cancer and head and neck cancer, respectively. MSI was used by 87% of the respondents for gastrointestinal cancer and 57% of the respondents for NSCLC. Oncologists reported that they were more likely to order each biomarker for cancers for which they perceive it as being more predictive, and less likely to use those biomarkers for cancers for which they perceive them as less useful in predicting the response (**Figure 3**).

**Figure 3.**
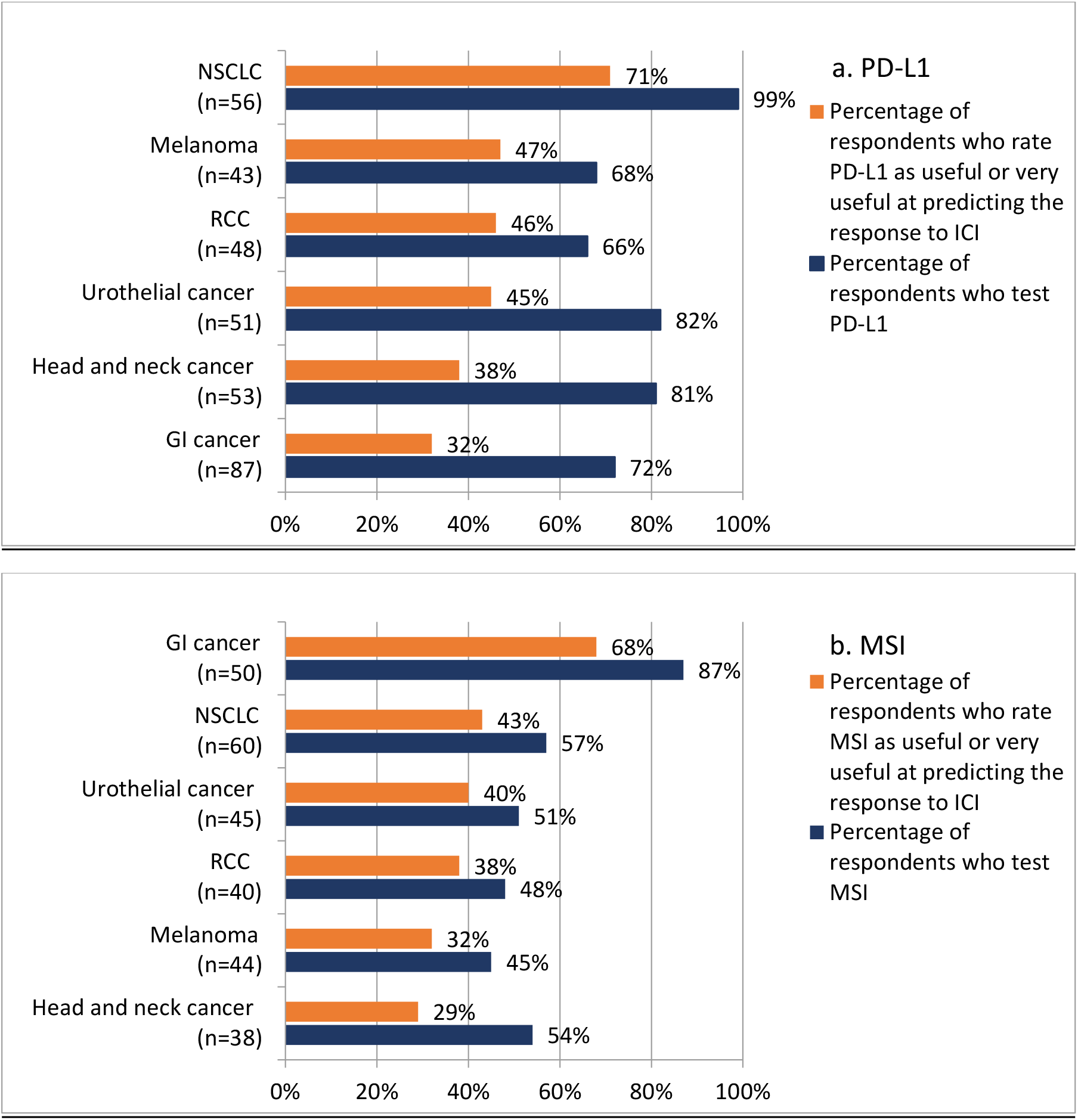
Association between the perceived usefulness of PD-L1 and MSI in predicting the response to ICI treatment and the use of these biomarkers in practice by type of solid cancer (n=100 respondents) Percentage of respondents who rated the usefulness of PD-L1 and MSI is based on scores on a 1 to 7 scale, with 1 being “not at all useful,” 6 being “useful,” and 7 being “very useful”

Notably, 22% of respondents reported not testing biomarkers for patients with RCC, 21% for patients with melanoma, and 11% and 9% for patients with head and neck cancers and bladder cancer, respectively. The top three reasons cited for not ordering a biomarker test were: lack of impact on clinical decision, patient refusal, and inadequate tissue.

About one-third of the respondents stated that they ordered biomarker testing when not required, with treatment planning being cited as the reason for doing so by half of that group.

More than half of the oncologists surveyed rated the NCCN guidelines, availability of published data demonstrating utility of the biomarker, and clinical indication or approval for the intended setting (i.e., drug label) as the most impactful factors for test ordering.

> “I do not like to waste resources especially if it does not change treatment course.” – verbatim comment by Oncologist, Community hospital

The need for more reliable prediction of treatment response was ranked by the survey respondents as the most prominent unmet need regarding biomarker testing for ICI treatment, with about one quarter of the respondents rating it as a 6 or a 7 on a 7-points scale (with a score of 7 being “very challenging”).

When asked in the follow-up survey about the required performance for a biomarker test to guide treatment decisions, 86% of respondents indicated that the highest acceptable rate of false negative test results (which would result in wrongly denying access to ICIs to patients who would indeed benefit from them) to guide their decision to not administer an ICI was 10% or less.

### Oncologists’ perception of a novel biomarker test

As part of both the interviews and the survey, oncologists were asked to comment on selected characteristics of biomarkers for predicting ICI response; specifically, by comparing a novel biomarker (based on peripheral blood glycoproteomic profiles) with currently available biomarkers (i.e., MSI, PD-L1, and TMB). Interview participants selected test performance, turnaround time, and being non-invasive as key attributes of a novel biomarker. Similarly, survey respondents rated high sensitivity as the most important requirement for adopting the new product in clinical practice, with less concern about limited specificity. Almost three-quarters of the oncologists surveyed reported that a test with a sensitivity of 90-95% would meet their needs (**Figure 4**). Most respondents indicated that the top reason for adopting a novel biomarker for predicting ICI response would be increased physician and patient confidence in deciding whether to prescribe ICIs (62% of respondents), in avoiding ineffective treatments (59%), and in decreasing unnecessary treatments and associated risk of irAEs (48%). Almost half of the oncologists queried indicated that they would start using the novel biomarker in their practice within three months of its availability, whereas one-third of the sample would start using it after three to six months. About 80% of respondents would use it to guide first-line treatment, whereas about 70% would use it for second- and third-line treatment.

**Figure 4.**
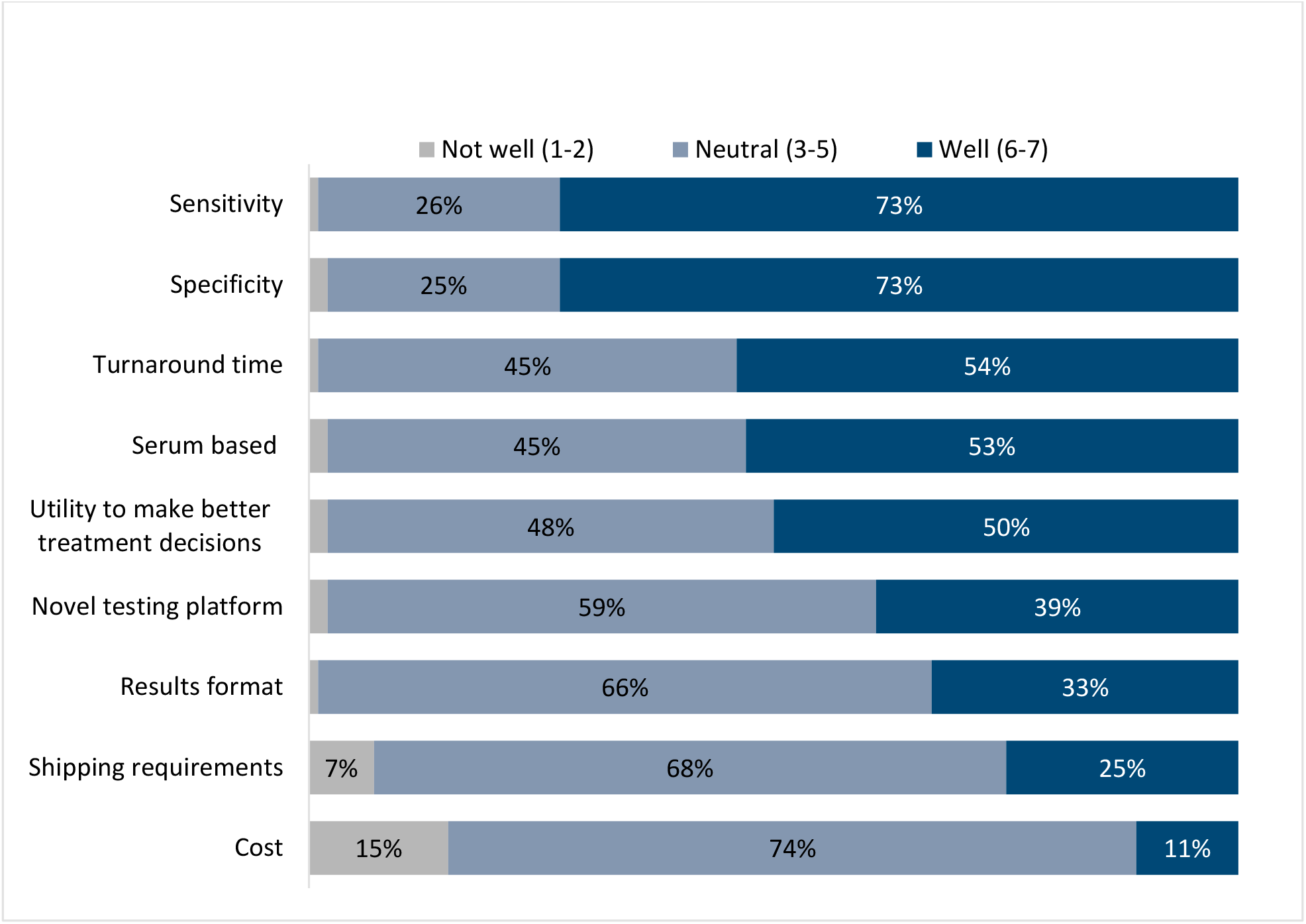
Characteristics of the new generation biomarker ranked according to oncologists’ perspective on how well they would meet treatment needs of patients with solid tumors (n= 100 respondents) Note: Bars corresponding to values ≤2% are not labeled.

## DISCUSSION

The approval of ICIs in 2011 has led to a change in the treatment paradigm for a range of malignancies^3,22^. Promising results from numerous clinical trials have generated enthusiasm for these drugs, which are currently approved for the treatment of a number of solid and hematologic malignancies^2^. Our study sought to explore oncologists’ perceptions on the clinical use of ICIs and related biomarkers. The study showed that oncologists have a very positive attitude toward use of ICIs in clinical practice and prescribe them for most patients who present with malignancies for which these drugs have been approved, especially as first-line combination- or mono-therapy. They indicated that cost is rarely a barrier for prescribing ICIs for approved indications and that physician preference is the primary decision factor. Oncologists highlighted the positive impact of ICIs on the care of patients with cancer, due to increased survival and reduced toxicity. The majority of surveyed oncologists indicated that the most important unmet need related to ICI treatment was the availability of more predictive biomarkers for treatment response. Clinicians indicated that such biomarkers would have great impact on their management of patients, reducing unnecessary treatments and side effects, as well as health care costs.

Noteworthy, compared to results in the published literature, oncologists surveyed generally overestimated the response rate to ICIs across most types of tumors^19,20^, which may result in prescribing these drugs more liberally than intended by the label, highlighting the importance of reliable therapy-response prediction biomarkers to avoid over-prescribing. This finding likely reflects a certain social desirability bias, resulting in selective recall of therapy success versus failure.

The availability of a testing platform that predicts more reliably likely responder or non-responder-status to ICI therapy than currently available genomics- or histology-based platforms, particularly one that does not require tissue, but can be run on peripheral blood samples has, for years, been elusive. Using a novel, vastly more informative class of analytes, i.e., post-translational protein modifications, this has now become possible. While the technology for the necessary resolution, namely mass spectrometry, has existed for many decades, managing the resulting very large and extremely complex raw data files has been prohibitive until very recently, when artificial intelligence and recurrent neural networks were applied to this task, resulting in the realization of the power of glycoproteomics and its potential for novel, highly accurate analytical assays.

Our study has certain limitations, many of which are common to studies employing the type of data and the mode of data collection utilized. First, given that the study participants agreed to be part of an online market research panel, it is possible that this resulted in a certain ascertainment bias, with answers not fully representative of a more diverse sample of clinicians, thus limiting the generalizability of the findings. To mitigate this limitation, the recruitment strategy was designed to enroll a mix of clinicians with academic and community practice affiliations. Second, given the self-reported nature of the information collected, observer bias is a possibility; to mitigate this risk, the following measures were put in place to: only a broad description of the study purpose was provided to the participants, participants were informed that data was collected anonymously and reported only in aggregate format, and industry standard-measures were used to ensure the quality of the data collected. The latter included: review for question straight lining, logical open ends, and data consistency, respondent verification process (validating identity and specialty, collecting National Provider Identified (NPI) and practice information), and duplicate respondent checks. Third, the research was sponsored by the company that is developing the novel test for predicting response to ICI treatment and the favorable attributes, in comparison with currently available tests, were presented to the respondents. To address this, the sponsor of the study was blinded to the study participants, and the data collection and analysis was completed fully independently by a third party.

## CONCLUSIONS

We believe that our study provides valuable information on oncologists’ perceptions and attitudes regarding ICI treatment and the current status of therapy-outcome prediction biomarker testing, as well as on the unmet needs in this important therapeutic area.

## Data Availability

All data produced in the present study are available upon reasonable request to the authors

## Author contributions

KL, PA, and MS designed the study and developed the study materials. All authors provided input into the data analyses, contributed to writing the manuscript, and read and approved the final manuscript.

## Acknowledgements

We are grateful to the oncologists who completed the interviews and surveys and to the researchers and staff at Olson Research Group who conducted the study.

## ABBREVIATIONS

AUC: area under the receiver operating characteristic curve
CTLA-4: cytotoxic T-lymphocyte-associated antigen-4
GI: gastrointestinal
ICI: immune-checkpoint inhibitor
KOL: key opinion leader
MSI: microsatellite instability
NSCLC: non-small-cell lung cancer
PD-1: programmed cell death protein 1
PD-L1: programmed cell death ligand-1
RCC: renal cell carcinoma
TMB: tumor mutational burden
NCCN: National Comprehensive Care Network
NCI: National Cancer Institute
SD: standard deviation.

